# Use of machine learning for triage and transfer of ICU patients in the Covid-19 pandemic period: Scope Review

**DOI:** 10.1101/2023.02.08.23285446

**Authors:** Lia Da Graça, Lucio Padrini, Richarlisson Moraes, Anacleta Rodrigues, Hugo Fernandes, Alexandre Barbosa de Lima, Monica Taminato

## Abstract

**Objective:** To map, summarize and analyze the available studies on the use of artificial intelligence, for both triage and transfer of patients in intensive care units in situations of bed shortage crisis so that health teams and organizations make decisions based on updated technological tools of triage and transfer.

**Methods:** Scope review made in the databases Pubmed, Embase, Web of Science, CINAHL, Cochrane, LILACS, Scielo, IEEE, ACM and the novel Rayyan Covid database were searched. Supplementary studies were searched in the references of the identified primary studies. The time restriction is from 2020, and there was no language restriction. All articles aiming at the use of machine learning within the field of artificial intelligence in healthcare were included, as well as studies using data analysis for triage and reallocation of elective patients to ICU vacancies within the specific context of crises, pandemics, and Covid-19 outbreak. Studies involving readmission of patients were excluded.

**Results:** The results excluded specific triage such as oncological patients, emergency room, telemedicine and non structured data.

**Conclusion:** Machine learning can help ICU triage, bed management and patient transfer with the use of artificial intelligence in situations of crisis and outbreaks.

**Descriptors:** Artificial Intelligence. Machine learning. Intensive Care Units. Triage. Patient Transfer. COVID-19.

## Introduction

Severe acute ICU syndrome coronavirus 2 (SARS-CoV-2) has infected millions of people around the world. In Brazil, from 3 January 2020 to November 25, 2022, there have been 35,104,673 confirmed cases of COVID-19 with 689,341 deaths^1,2^. These figures represent 10.3% of total deaths in the world. A specific academic work published data about death toll velocity and acceleration of contamination of Covid-19. It presented the result that “the most important factor is the demand for critical care beds”^3^. Consequently, the availability of healthcare resources such as ICU beds resulted in deceleration of the mortality rate in the current Sars-COV-2 pandemic crisis in countries such as Italy and Spain^4^.

In the current pandemic experience of managing *COVID-19* disease the extent of the impact of the disease on people and health services challenged existing resources. We observed that countries were not prepared for such a response or did not have sufficient resources to manage this crisis, presenting insufficient emergency response in the crisis^5^. Among the relevant aspects to face the pandemic, the availability of beds is considered a causal variable of increased mortality rate by COVID in Brazil^6^. Maximization of health resources and speed of response to health crises affect both developed and developing countries, western and eastern countries, from both hemispheres, indistinctly. In places such as in the Lombardy region (Italy), where intensive care beds do not absorb all the demand of patients, and professionals have been placed in a dilemma of who will take the bed^7^.

In other cases, as in Brazil, system overload can be a problem in the next epidemic or endemic crisis. This circumstance of difficult patient allocation - especially in ICU beds - arises when after all hospital administration movements or resource transfers have already been made, and yet resources remain limited. The limited number of hospital beds forces ICUs to change their triage approaches, as there is pressure for intensive use of resources, while still monitoring the patient adequately. And as expected, some studies have shown that decreased ICU bed availability is related to a lower likelihood of ICU admission^6.^ Hospitals avoid acquiring more beds due to higher fixed maintenance costs and potential waste of resources when these beds are not used.

In this context, searching for an ICU triage model through resource optimization criteria becomes important, in order to facilitate the access of patients to treatments in a convenient time and to guide health professionals. The objective of this scope review is to identify and analyze the contributions and advances of artificial intelligence (AI) in the ICU bed triage work and transfer of patients due to the lack of ICU beds in order to guide physicians to make decisions based on technological advances.

## Material and methods

### Study design

Background and research question: Evidence of the need for ICU beds and subsequent transfer of patients in the absence of beds. We aimed to find works in which health teams, challenged by the shortage of ICU beds in crisis situations, can count on the aid of artificial intelligence tools, which gives rise to the research question: are there currently algorithms capable of simultaneously providing a intelligent quick triage and subsequent transfer of the patient to another health unit, in case of unavailability of an ICU bed?

This Scoping Review study followed the recommendations proposed by Arksey and O’Malley^7^ and the Jonna Briggs Institute (JBI)^8^. It allows mapping the main concepts and indicators, research areas and identifying knowledge gaps. For the construction of the research question, the Population, Concept and Context strategy was used (P: Patient, C: Intensive Care Unit - ICU, C: capacity of beds, availability of drugs, equipment and intensivist team) for a scoping review.

This was a scoping review developed by researchers of the CNPq research group on Epidemiology, Systematic Review, and Health Policies of the Paulista Nursing School, Polytechnic School of the University of São Paulo and the Naval War College’s Simulations and Scenarios Laboratory. The review was registered on Open Science Framework in October, 2022 (https://osf.io/wyvgd//). In addition, the reporting of this Scoping Review followed the recommendations of the PRISMA for Reviews (PRISMA-ScR)^9^.

### Search of relevant studies in the databases

Searches were conducted using medical subject headings in the following databases: Pubmed, Embase (Elsevier), Web of Science, Cumulative Index to Nursing and Allied Health Literature - CINAHL, Cochrane, Latin American and Caribbean Health Sciences Literature - LILACS, Scopus, Scielo, Institute of Electrical and Electronics Engineers - IEEE, ACM Digital Library. Additionally, the Rayyan platform offered a selection of studies on Covid-19 keyword which we included in the search. A manual search was conducted using the references for the primary and secondary studies found in the electronic search. The Search strategy was developed in Pubmed/MEDLINE and adapted and used for each electronic database, according to the particularities. Searches were conducted in October 2022 except for the search in IEEE which was made in December 2022. There were no restrictions on languages or forms of publication.

### Selection of studies

Identification of eligible studies followed a two-stage process accomplished by two independent reviewers. Any disagreement was resolved by reaching a consensus. In the first stage, after exclusion of duplications, the titles and absIntelligents of the references identified through the search strategy were evaluated and the potentially eligible studies were pre-selected. In the second stage, a full-text evaluation of the pre-selected studies was carried out to confirm their eligibility. The selection process was performed through the Rayyan platform (https://rayyan.qcri.org).

A manual search was conducted in the references of the primary and secondary studies that were identified through the electronic search. The search strategies developed and used for each electronic database were performed in October, 2022. They are presented in Table 1.

**Table 1.** Search strategies developed and used for each electronic database (October, 2022; December, 2022).

| ELECTRONIC DATABASE | Number of articles found | SEARCH STRATEGY |
| --- | --- | --- |
| Pubmed | 421 | ((((((("ICU Triage"[Title/AbsIntelligent]) OR "ICU Screening"[Title/AbsIntelligent]) OR Patient Transfer [Title/AbsIntelligent]) OR intelligent triage [Title/AbsIntelligent]) OR ICU algorithm[Title/AbsIntelligent]) OR "ICU automatic triage"[Title/AbsIntelligent]) OR ICU machine learning[MeSH Terms])) AND (((((covid-19[MeSH Terms]) OR SARS-CoV-2[MeSH Terms]) OR ICU Transfer,[MeSH Terms]) OR Intensive Care Transfer [Title/AbsIntelligent])) |
| Embase | 168 | #1 'ICU triage'/exp OR (ICU screening) OR (intelligent triage) OR (automatic triage) OR (ICU transfer) OR (patient transfer) OR (intensive care transfer) OR (algorithm triage) OR (intelligent transfer) OR (critical patient transfer) AND #2 'machine learning'/exp OR artificial intelligence OR (algorithm) OR 'automatic transfer'/exp OR triage protocol * AND #3 'Sars-Cov-2'/exp OR 'Covid-19'/exp |
| Web of Science | 203 | ((("ICU intelligent triage" OR "ICU automatic screening" OR transfer* OR ICU severity grade* OR "ICU triage protocol") AND ((machine learning AND artificial intelligence AND "algorithm") OR Covid-19 ICU OR "Sars-Cov-2 ICU" OR ICU patient transfer)) |
| CINAHL | 40 | #1 (ICU Triage) OR (Screening, Critical Care) OR (Automatic Triage, Intelligent Triage) OR (Automatic Screening) OR (ICU protocol, intensive care) OR (ICU transfer, patient transfer) AND #2 (Machine Learning) OR (Artificial Intelligence, Algorithm) OR (Data Mining, Automatic Tool) OR (ICU transfer, Triage) OR (Intensive Care, Artificial Intelligence) OR (Machine Learning, ICU transfer) OR (Covid-19) OR (Sars-Cov-2) OR (Critical Care) or (Patient Transfer) OR (Algorithm) OR (Intensive Care Unit transfer) OR (ICU Screening) OR (ICU automatic screening) OR (ICU intelligent transfer) OR (Machine Learning, Tool) OR (Artificial Intelligence, Tool) OR (Artificial Intelligence, Patient Transfer, Machine Learning) OR (ICU Transfer, Triage) OR ICU* |
| Cochrane | 9 | ID Search #1 MeSH descriptor: [ICU Intelligent Triage] explode all trees #2 (ICU, Intelligent Triage) or (Intelligent, ICU Triage,) or (Intelligent Triage, ICU) or (Triage, ICU Intelligent) or (Triage, Intelligent ICU) #3 #1 or #2 #4 MeSH descriptor: [ICU machine learning Screening] explode all trees #5 (ICU, machine learning, screening) or (ICU, screening, machine learning) or (machine learning, ICU, screening) or (machine learning, screening, ICU) or (machine learning, ICU, screening) or (screening, ICU, machine learning) or (screening, machine learning, ICU) or (ICU transfer) or (Critical patients transfer) or (ICU transfer) or (patient transfer) or (algorithm) or (transfer algorithm) or (triage algorithm) or (ICU algorithm) or (screening algorithm) or (intensive care unit, algorithm) or (intensive care, algorithm) #6 #4 or #5 #7 MeSH descriptor: [Intensive Care] explode all trees #8 (Mask) #9 #7 or #8 #10 #6 or #9 #11 MeSH descriptor: [ICU Hospitalization] explode all trees #12 (ICU Hospitalizations, transfer) or (ICU Hospitalized, Transfer) or |
|  |  | (ICU hospitals, Transfer) or (Intensive Care Transfer) or (ICU, Transfer) or (Intensive Care Transfer) or (Patient, Transfer) or (Critical Patient, Transfer) or (ICU Patient, Transfer) #13 #11 or #12 #14 MeSH descriptor: [ICU life expectancy] explode all trees #15 (ICU, life expectancy) or (algorithm, ICU Life Expectancy) or (ICU life expectancy, transfer) or (transfer algorithm, ICU life expectancy) or (machine Learning, ICU life expectancy) or (Transfer Algorithm) or (Hospital Transfer) or (Hospitalization, Transfer) or (Hospitalized, transfer) #16 #14 or #15 #17 #13 or #16 #18 #3 AND #16 AND #17 |
| LILACS | 1 | (Covid-19 OR SARS-CoV-2) AND (Algorithms OR artificial Intelligence OR machine learning) AND (unit intensive care OR Intensive Care Units OR emergency) AND (screen* OR triage) |
| Scielo | 0 | ("ICU triage" OR "Intensive Care Unit Triage " OR Screening* OR Intelligent Triage* OR "Automatic Triage" OR Patient Transfer* OR ICU Transfer* OR "Transferência de UTI" or "Triagem de UTI") AND ((Machine Learning AND Artificial Intelligence AND "Covid-19") OR Data Mining OR Sars-CoV-2 "Triagem inteligente de UTI" OR Transferência de Pacientes de UTI OR Algoritmos) |
| IEEE | 33 | "All Metadata": "covid 19" OR "All Metadata": "SarCOV 2" OR "All Metadata": "nCoV" OR "All Metadata": "2019nCoV" OR "All Metadata": "19nCoV" OR "All Metadata": "COVID19" OR "All Metadata": "COVID" OR "All Metadata": "SARS-COV-2" OR "All Metadata": "SARSCOV-2" OR "All Metadata": "SARS-COV2" OR "All Metadata": "SARSCOV2" OR "All Metadata": "SARS coronavirus 2" OR "All Metadata": "Severe Acute Respiratory Syndrome Coronavirus 2" OR "All Metadata": "Severe Acute Respiratory Syndrome Corona Virus 2") AND ("All Metadata": "artificial intelligence" OR "All Metadata": "machine learning" OR "All Metadata": Algorithms) patient transfer triage OR screen* |
| ACM | 43 | (Covid-19 OR SARS-CoV-2) AND (Algorithms OR artificial Intelligence OR machine learning) AND (unit intensive care OR Intensive Care Units OR emergency) AND (screen* OR triage) |
| Rayyan | 13 | Covid-19 Database: New Review, Other Reviews, Covid-19 Dataset, Show, Rayyans's NPL-enabled search, literature review, copy. |

Source: Table prepared by the authors using information provided by the database queries.

We used as eligibility criteria the presence of content on Covid-19 ICU intelligent triage in any part of the world, their application and the use of machine learning (ML) or similar uses. Types of Interventions Studies involving ICU triage, ICU screening, transfer of patients, ICU hospitalization, ICU life expectancy, artificial intelligence, machine learning, ICU bed occupancy were considered eligible. Considering the limited number of studies to evaluate the theme the objective of this brief review is to map existing knowledge on the subject and identify study designs by the level of evidence.

## Results

The electronic search strategies retrieved 931 references. Duplicate articles numbered 362, which were eliminated. The researchers’ elimination criteria for eliminating references from the remaining 573 papers were: use unstructured data, specific ICU screening cases like cardiac and oncologic, systematic review articles, scope reviews, ICU readmission, emergency room triages, telemedicine, use of mobile phone data. The filters eliminated 457 works. There were 33 titles in the “conflict” classification, with tie-breaking done by the second researcher in the second stage (blind off). Of these in conflict status, 32 papers were eliminated for criteria that may appear in more than one article. Exclusion also occurred because the same article was presenting more than one reason for exclusion.

The exclusion criteria in maybe status were: dealing solely with mortality prediction, not dealing with ICU setting, use of imaging only, response to Covid-19 infection, responses to the pandemic, pandemic management in municipalities, Covid-19 symptom detection, strategies for the general population, Covid-19 infection case detection, general use of digital tools, screening for respiratory problems, frontline policy, hospital admissions. Two independent reviewers selected the studies and discrepancies were resolved by three senior reviewers. Reading the text of the 80 selected references, only 7 studies remained eligible. We present the flow chart of the selection process in figure 1.

**Figure 1.**
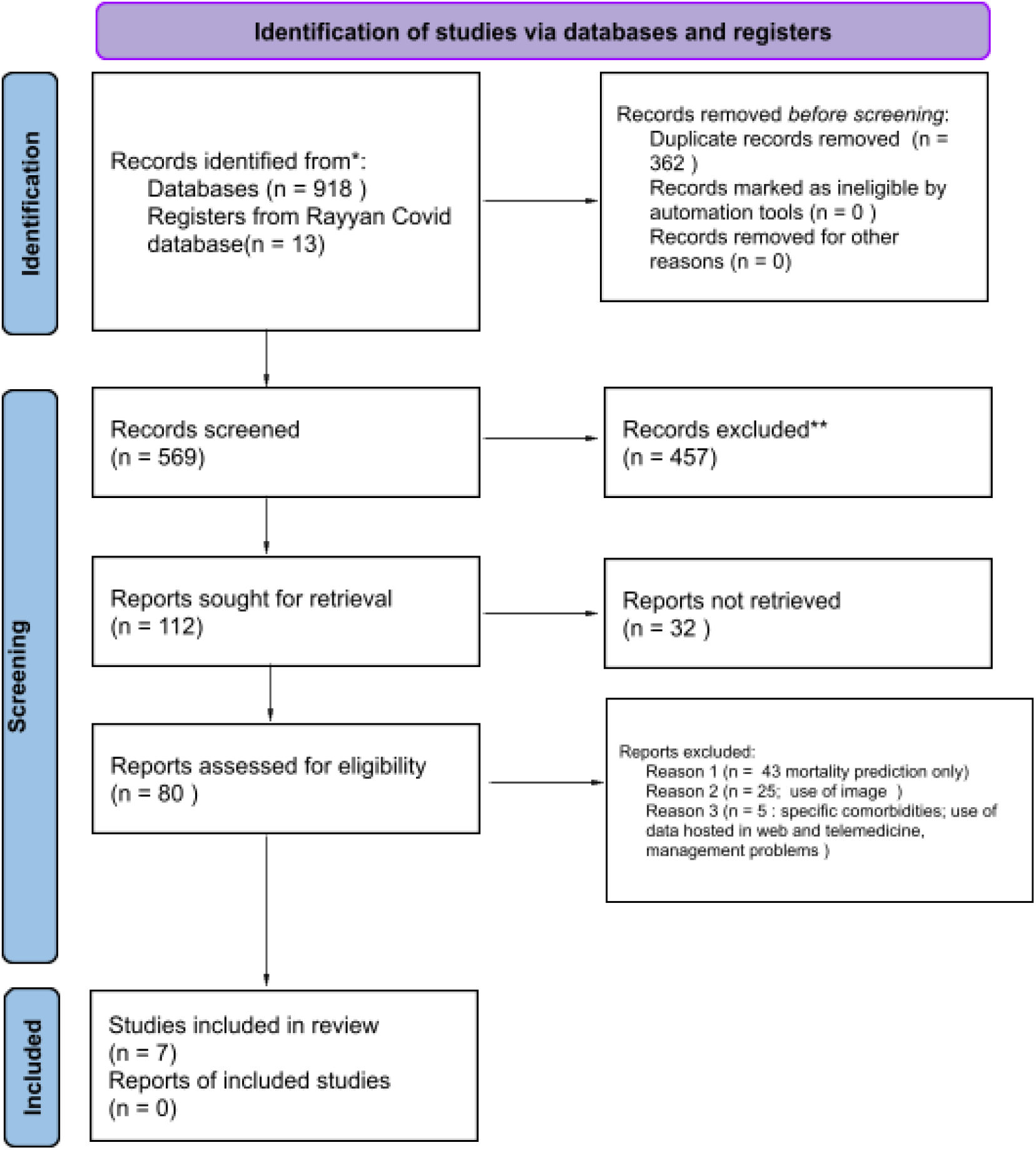
Articles selection PRISMA 2020 flowchart diagram for new reviews. elaborated by the authors from the original PRISMA: Page MJ, McKenzie JE, Bossuyt PM, Boutron I, Hoffmann TC, Mulrow CD, et al. The PRISMA 2020 statement: an updated guideline for reporting systematic reviews. BMJ 2021;372:n71. doi: 10.1136/bmj.n71. For more information, visit: http://www.prisma-statement.org/. At this stage, the blind on button of Rayyan application was disabled, so that the articles in maybe and conflicting statuses could be resolved.

### Characteristics of the selected articles

As for the timeliness of the selected works, the two oldest are from January and June 2020 and five were published in 2021.

The study from the United States^10^ describes screening strategies for COVID-19 patients and risk stratification for the need for an ICU bed12. The purpose of the study was to develop an assessment for all patients at high risk of clinical deterioration, using a risk prioritization tool. Such a tool used ML and a 24-hour ICU transfer forecast, seeking to facilitate efficient use of care providers’ efforts and help hospitals plan their flow of operations. Instead of placing patients in a priority queue due to the immediate need for an ICU, the researchers assessed the need for ICU beds, by predicting clinical-medical deterioration, using the retrospective cohort method, composed of admissions due to COVID-19. The data collection period was between February 26 and April 18, 2020. The input temporal data series includes vital signs, nursing staff assessment, clinical laboratory data, and electrocardiograms.

Wiesner et al.^11^, developed a study focusing on guaranteeing ICU vacancies, through the predictability of discharge and transfer of their patients. It used as parameters: a) establishment of a communication and coordination structure under the direction of a specialized center for COVID-19 cases; b) joint clinical assessment of disease severity and an assessment of current capacities at the site and in the network, the coordinating center determines the patient’s further whereabouts: in a COVID-19 ICU or at another center; c) allocation capacity in ICU and geographic locations, clinics with intensive care units were identified by a group of experts who, together and in conjunction with the coordination center, should treat patients seriously ill with COVID-19. Telemedicine was the basis of this coordinated network, with daily bedside meetings and consultations between the coordination center and the other COVID-19 ICUs possible. The flowchart of the algorithm called “POST-SAVE” developed structures the additional care of patients with long-term ventilation (patients ventilated invasively, tracheotomy or ventilated non-invasively) to guarantee intensive care capabilities in clinics, when they are in a situation of acute resources sold out. There is a transfer to ventilation weaning units. The first step is the central recording of POST-SAVE ventilatory capacities (clinical and non-clinical ventilation facilities). The allocation and transfer of patients from intensive care units can thus be guaranteed. The weaning potential is checked in clinics. All patients to be transferred are categorized into 3 groups according to clinical criteria in groups: Group 1: Patients with high potential for immediate weaning; Group 2: Patients with low immediate weaning potential; Group 3: Patients without potential (actual) weaning. The results presented were the algorithms 1) of daily screening of the respiratory situation during weaning and 2) of daily screening of the respiratory situation during weaning. Research also includes the use of a standardized digital database, the use of trained controllers to facilitate communication between intensive care units and weaning units, and the establishment of a prospective data register to be able to reassess patients for its weaning potential.

The study by Crowley et al.^12^ was developed by an American team and aimed to verify the risk factors of patients with COVID-19 with indications for ICU admission. Biomarkers, clinical risk factors, and comorbidities were used to predict/track the clinical outcomes of 5,568 patients, in a retrospective medical record review of patient admissions, which occurred between March 1 and May 1, 2020, with patients older than 18 years, for positive Covid-19 in less than 15 days before admission.

The researchers from Italy^13^ developed a prognostic model in ML for the progression of COVID-19. The researchers addressed the prediction of intensive care unit (ICU) admission within 5 days. Three ML models were developed based on 4,995 complete blood count exams. Three different models were proposed: two fully interpretable models and a black-box model. This model was obtained by determining the best combination between five different classes of ML models: gradient augmentation^1^, logistic regression, support vector machine (SVM), RF and a decision tree. The researchers concluded that blood count data and ML learning methods can be used for the economic prediction of ICU admission of patients with COVID-19. In particular, they demonstrated how the blood count collection through routine blood tests can be used for ML models, in situations of limited daily screening resources. The researchers intend to validate their models externally, with data from other hospitals and other periods of time, including different treatments and therapies applied to patients^15^.

The research developed by Deif et al.^14^ was developed with collaboration of members from different countries - Egypt, Turkey, Korea and Thailand. Comparing algorithmic methods, they developed a classifier integration system called Xtreme Gradient Boosting (XG Boost). The system seeks to assist health authorities in identifying the priorities of patients to be admitted to ICUs according to the results of the biological laboratory investigation for patients with COVID-19. The XGBoost classifier was used to decide whether or not they should admit patients to ICUs, before applying it to an *Analytic Hierarchy Process* (AHP) to rank priority of ICU admissions. A total of 38 clinical variables were used. In this research, five types of classifier algorithms were compared by running the XGBoost classifier: Support Vector Machine (SVM), Decision Tree (DT), K-Nearest Neighborhood (KNN), RF and Artificial Neural Network (ANN), to evaluate the performance of XGBoost. In parallel, the proposed Analytic Hierarchy Process (AHP) classification compared the results with a committee consisting of experienced physicians.

The study by Vagliano et al.^15^, proposes automated machine learning *design* (AutoML), which includes automatic selection of ML models. Manual variable selection was discarded as restrictive, inefficient, and biased. Dataset contained demographic data, minimum and maximum values of physiological data in the first 24 hours of ICU admission, diagnoses (reason for admission as well as comorbidities), ICU, as well as data on in-hospital mortality and length of stay. The analyses were run on two different sets: 1) including only variables available at ICU admission and 2) all variables available after the first 24 hours of ICU admission. The research evaluated the performance of AutoML for modeling clinical prognosis by comparing classical modeling (manual selection of variables followed by regression) and AutoML modeling approaches. In particular, the performance of Auto Prognosis, a proposal for a tool developed for modeling clinical prognosis that simultaneously learns 20 ML models (for example, regression, neural networks and linear discriminant analysis) for the prediction of intra-hospital mortality, was evaluated. hospitalization of patients with COVID-19 who were admitted to the ICU. In the case of new diseases, the selection of predictors is not possible.

The third study from USA^16^ calls attention to the fact that their ML models were more consistent in predicting mortality than ICU admission meaning that ICU admission decisions are more variable “because of how each physician practices and of how the pandemic has progressed temporally”. The authors advise researchers must find themselves what clinical variables are the best in order to predict mortality in the ICU triage, for the fact there is no consensus about that. Additionally, only a few academic papers worked on the Covid-19 mortality prediction and its associated variables using logistic regression with a limited number of variables^19^. The authors encourage the use of ML is increasingly being used in medicine, because of its ability to analyze a large number of variables^20^. Nevertheless, the authors advise replication of tests in other health institutions and countries and alert the fact that the variables in their dataset were collected in the admission of the patient at the admission of the patient in one of the pandemic peaks. The results show different ML algorithms which are able to predict the need for ICU admission, or internal transfer from hospitalization or emergency department to the ICU unit. It does not project the transfer of the patient to another hospital.

## Discussion

The first contribution of this review is to map the triage criteria for crisis situations and their potential removal and transfer. This allows systems and physicians to prepare in advance for a large influx of patients. Accurate triage systems and appropriate allocation of limited resources will be essential components in ongoing response to ongoing challenges, quickly identifying patients who require immediate attention, optimizing the efficient use of medical resources. Additionally, observing the heterogeneity of criteria for a patient’s admission to the intensive care center can assist nursing and healthcare professionals in saving time and aid in decreasing selection biases. The goal of a triage system is to direct limited resources to those patients most likely to benefit from them. Implementing a screening system requires careful coordination between health managers, nurses, physical therapists, physicians, health systems, local and regional governments. The greatest possible transparency with the public is advised, with the goal of maintaining trust. Triage is the course of action we take when we have exhausted our capacity to expand our acute care resources. Surge capacity refers to the ability of a hospital or other health system to expand its normal operational capacity in the setting of an emergency^20^.

Improving the workflow for ICU bed triage starts from protocol analysis and triage decision to the point of being able to reduce as much as possible (and if possible zero) the delay between the immediate request for a bed opening until it is obtained. An ICU bed opening can be tracked by communication between health care units, enabling patient transfer. Specifically in Brazil, data on bed requests can be observed in the Central Regulation of Health Service Offers (CROSS), created in 2016. The Public Health Management System CROSS seeks to meet the prerogative called “zero vacancy” which in Brazil was set forth in Ordinance No. 337 of March 24, 2020^21^. This regulation provides immediate access to assistance to patients at risk of death or intense suffering. In this case of research, the availability of an ICU bed should be considered an exceptional situation and not a daily practice in emergency care.

Among the studies found which evaluated the screening criteria, all those that used images were excluded, because they deal with data that require more time and complexity of analysis, which can compromise the speed of a screening workflow. The focus was on the type of clinical-medical variables considered as databases for analysis, and which ML tools performed best. In addition to statistical techniques, there are a myriad of ML tools for data analysis. Of these, a few were explored by the researchers in the seven chosen studies. In one of them, the algorithm does not make explicit which tool was used, probably because it is a contractual clause from the Berlin city government to use the algorithm^11^. This study was the only one that includes patient transfer planning. However, it did not try to classify by priority of mortality of the patient, but by need for invasive ventilation.

Intensive care professionals may have doubts about the implications of triage in a disaster situation (accidents, war or epidemics) because of a lack of experience in the context, combined with a tendency to confuse it with triage that is done in routine hospital emergency rooms. The difference is that in disasters, in addition to sorting and prioritizing patients, triage also includes the optimization of scarce resources, which implies the focus of resource management administration on individual patient outcomes at the population level. There are many practitioners with professional experience in prioritizing patients. However, few clinicians have experience in making population-level decisions during periods of scarce resources^20^. This concept of triage in crisis situations is discussed by Christian^22^ and by Sprung et al. ^23^. The first author explains that both impact on the individual’s chance of survival (e.g. delayed treatment to individual patients) and inappropriate use of resources (personnel, equipment) can hinder the proper outcome of population triage. Screening in this context involves greater complexity, and not a binary outcome between death and survival, but rather the probability of death across a range of treatment options. Thus, the key factors being considered in making disaster triage decisions must include survival and the resources needed to achieve these outcomes in specific situations.

The problem of patient ICU triage and subsequent transfer from ICU units (caused by ICU bed shortage) this contingency was not discussed in the algorithms we found. There are some gaps in the literature such as how AI can assist less experienced healthcare professionals who may deal with shortage of resources to deliver adequate care through learning from previous experiences ^35-36^. Additionally, some AI applications showed results in medicine with high accuracy, sensitivity and specificity using different models and algorithms^38-39^. As we have observed during this revision, the use of AI is expected to increase in different health domains^40-41^. In brief, researches of this interdisciplinary field are motivated by a) the possibility to analyze big datasets of different types of variables; b) the use of precision predictive analytics c) optimization of the use of resources; d) improvement of medical and nursing clinical nursing decision-making process; e) the unexplored potential for medical and nursing automated systems implementation^42-43^.

Study limitations are: a) exclusion of studies on artificial intelligence tools that do not use structured data; 2) only the study led by Egypt used Brazilian medical clinical data from Hospital Sírio Libanês and 3) limitation of the type of crisis: pandemic between 2020 and 2022, which does not include other types of emergencies such as floods, fires and war.

## Conclusion

No papers were found on customizable solutions of the combination triage and transfer of ICU patients or the transfer of the patient to a remotely available ICU vacancy, after the triage by severity classification of the patient, made by a tool developed through ML. The use of automatic protocols and fast decisions can be considered a necessary response to the aftermath of a disaster or pandemic. Thus, intelligent and automated or semi-automated triage and transfer and indication of the need for ICU beds to central management (private or public) allows the entire workflow of healthcare professionals and intensive care management to better assist patients. It is expected that the development of automated tools in this regard will represent the availability of ICU beds in an organized way, at an appropriate time for patients and health care professionals management.

The COVID-19 pandemic remind us of previous lessons about responses to accidental cases with multiple victims such as vehicle accidents, victims of natural disasters such as earthquakes, floods, tornadoes, tsunamis; casualties in wars and conflicts, industrial disasters such as fires, explosions, toxic spills, etc.; and the historic epidemics of Spanish flu, AIDS and H1N1. The published works are focused on specific problems generated by the needs in the pandemic period. To obtain an intelligent ICU triage model in high generalization that provides patient transfer, it is necessary to know what automatic protocols already exist, what variables were used in the data analysis and what are the adequate technological tools available. In addition, it is necessary to describe patient classification decisions and their tie-breaking criteria, reducing decision asymmetries between health teams. Accordingly, development and implementation of a triage and transfer algorithm will enable careful coordination between health managers, nurses, physicians, physical therapists, health care facilities, and governmental spheres.

## Data Availability

All data produced in the present study are available upon reasonable request to the authors

## Footnotes

1 Gradient-based Data Augmentation (GDA) is calculated from the image pixel value gradient of the posterior probability distribution that is the model output^19^.

